# Daily Salt Intake, its Discretionary Use and Validation of Methods for Estimation using Spot Urine – Findings from Islamabad, Pakistan

**DOI:** 10.1101/2020.07.09.20149468

**Authors:** Muhammad Arif Nadeem Saqib, Ibrar Rafique, Muhammad Ansar, Tayyaba Rahat

**Author notes:** **Corresponding author:** Ibrar Rafique, Research Officer, Pakistan Health Research Council, Shahrah-e-Jamhuriat, Constitution Avenue, G-5/2, Islamabad.

## Abstract

**Background:** The study was designed to estimate daily salt intake, its discretionary use in healthy individuals and to validate three common methods for salt estimation in Pakistani population.

**Methods:** Information on demography and discretionary salt use was collected healthy adults (>18 years) along with a blood sample, spot urine sample and 24 hours urine samples. Sodium, chloride, potassium levels and serum creatinine were measured using standard methods. For daily salt estimation, three common methods i.e. INTERSALT, Tanaka and Kawasaki were validated for their applicability in local settings.

**Results:** Overall 24 h sodium excretion was 158 mmol/l indicating intake of 8.64 (±4.43) grams salt per day which was significantly associated with male gender (p. <0.004) and adding salt during cooking (p. <0.0001). Most (73%) of the participants know about hazardous effects of high salt intake, however, only 25% consider important to lower salt intake. Although, there is insignificant difference (p. 0.09) between measured and estimated 24 hour urine however none of three methods i.e. INTERSALT(bias: -19.64; CCC -0.79), Tanaka(bias: 167.35; CCC -0.37) and Kawasaki (bias: -42.49, CCC -0.79) showed any agreement between measured and estimated 24 hour sodium.

**Conclusion:** Daily intake of salt was high than recommended by the WHO. Findings showed that none of the three methods could be used for estimating daily intake of salt in local settings of Pakistan.

## Introduction

High intake of salt is associated with increased risk of hypertension, heart diseases and gastric cancer (1–3). In most of the countries, average daily salt intake ranges 9-12□g/day that is much higher than recommended i.e.<5g/day(4). It has been reported that non-communicable diseases (NCDs) burden could be decreased in the population by lowering salt intake(5). This will help in reducing the burden of NCDs globally with low cost. (6). Keeping in view the importance, the member states of WHO agreed to usefulness of reducing salt intake, the WHO member states have agreed for 30% decrease in sodium intake till 2025(7).

Estimation of daily salt is an important step to observe the level of sodium intake among general population which can provide the way forward to public health initiatives. intake (8). For daily salt estimation, the 24 hour urine collection is the standard method for salt estimation but is laborious and troublesome especially for healthy individuals(9). In order to overcome hurdles associated with 24hours urine collection, different mathematical equations have been formulated using spot urine which is being used as a appropriate. (10–15). Although spot test does not provide accurate prediction for 24hours sodium, however, this test can be used for population estimates and can provide adequate information for monitoring(13, 16).

Despite the rising burden of NCDs, especially hypertension and Cardiovascular diseases (CVDs) in Pakistan (17), information on the daily salt intake and its discretionary use was not available. Similarly, to our best knowledge, there was no report available addressing the issue of daily salt estimation in the local population. Therefore, this study was planned for estimation of daily salt intake we planned this study to estimate the daily salt intake in healthy individuals of Islamabad along with their discretionary usage. Besides this, it was also assessed whether these three methods are applicable for estimation using spot urine.

## Methods

This was a descriptive cross-sectional study conducted at Islamabad. Total of 120 healthy subjects aged more than 18 years were enrolled using non-probability convenient sampling. Those who had a history of hypertension or currently hypertensive, taking any medications, difficulty in collecting urine and had pregnancy were excluded. It was assured that participants were healthy and were not suffering from fever, diarrhea or any other infections at the time of sample collection.

Information about demographics, discretionary salt use, awareness about the hazards of increased salt intake and their attitude for salt control was collected from the residents of Islamabad using a pre-designed questionnaire. Weight and height were measured as per standard protocol. The data was collected by a trained data collector who also briefed the participants about the process of sample collection. All testing was done at Excel Lab (Pvt) Ltd which is registered in the Programs for Excellence in Laboratory testing of the College of American Pathologists. The Excel Lab (Pvt) Ltd has sample collection points in the entire city of Islamabad.

After enrollment, data of each participant registered in Excel lab (Pvt) Ltd database along with detail of tests to be carried out which automatically generated a unique identification number along with barcodes for each test. These barcodes were placed spot and 24-hour urine collection containers and gel tube for a blood sample.

Two labeled collection containers (each separate for 24 hour urine and spot urine sample) were given to the participants. All participants were briefed about the process of collection of both spot and 24 h urine collection. They were asked to discard the first urine sample at 8.00 am on starting the day while including the first sample on the second day at 8.00 am.

The participants collected 24 hour urine samples as per convenience but preferably on the weekend. Majority of samples were collected on the weekend where the participants started their urine collection on Saturday morning till Sunday morning. The blood samples were collected during the urine collection for estimation of serum creatinine and electrolytes. The participants living in close vicinity of these collection points dropped their sample at their nearest point in the morning after completing 24 hour collection of urine. The participants provided second morning urine sample as a spot sample when coming to Laboratory. From these points, samples were immediately transported to main laboratory for testing.

Those participants, who lived far from collection points or reluctant to visit collection point, data collectors themselves visited their home in the early morning. They also provided second morning urine sample as a spot sample. This helped us to assure that second-morning sample was taken as spot sample and was not part of 24 hour urine sample. The data collectors picked samples from their home and directly dropped at the main lab for analysis.

Each 24 hour sample was assessed for its completeness. Those samples having volume <0.5 mL and reported missed collection were discarded. Similarly samples collected outside the range of 24±2h were also excluded. All samples were analyzed on the day of their collection. The potassium, sodium, and chloride were tested using electrode method while serum creatinine was done using the kinetic Jaffe reaction along with controls.

Institutional Bioethics Committee of [removed for blind peer review] provided ethical clearance. The research work carried according to the Declaration of Helsinki. The participants provided written informed consent prior to their participation.

### Statistical analysis

The data was entered in MS Excel and analyzed using SPSS 21 (SPSS & IBM, Inc, Chicago, Illinois, USA) for analysis. Tukey test (18) was used to check outliers at either side, categorized as ‘outside’ or ‘far out’ values. The outside and far out values were removed from data. Finally data of 106 participants was analyzed.

Gender wise difference between demographic characteristics and various analytes was determined using independent sample t-test while discretionary salt use and its association with risk factors was assessed by multivariate logistic regression.

The intake of salt on daily basis was calculated by the formula described elsewhere(14) while 24 hour sodium in urine was estimated using and Tanaka (10) Kawasaki (15), INTERSALT (19)methods (Table 1). Paired t-test was applied to assess the difference between 24 hour measured and estimated salt intake. Correlation methods (concordance correlation co-efficient and intraclass consistency was used for determining the validity of three methods for estimating 24 hour sodium by visualizing on scatter plot. Systematic bias between measured and estimated sodium was assessed by Bland and Altman plot. P-value (<0.05) was considered statistically significant.

**Table 1.** Equations of three methods used to stimated24-hours urinary sodium excretion using spot urine.

| Method | Urine Sample | Formula to Estimate 24-h urinary sodium excretion (mg/day) |
| --- | --- | --- |
| **INTERSALT** | **Casual Spot Urine** | $23 \times ((25.46 + 0.46 \times \text{Naspot}) - 2.75 \times \text{Crspot} - 0.13 \times \text{Kspot} + 4.10 \times \text{BMI} + 0.26 \times \text{Age})$ **(Male)**<br>$23 \times ((5.07 + 0.34 \times \text{Naspot}) - 2.16 \times \text{Crspot} - 0.09 \times \text{Kspot} + 2.39 \times \text{BMI} + 2.35 \times \text{Age} - 0.03 \times \text{Age}^2)$ **(Female)** |
| **Tanaka** | **Casual Spot Urine** | $23 \times 21.98 \times (\text{Na spot} / \text{Crspot} \times \text{PrUCr24h})^{0.392}$ $\text{PrUCr24h} = 14.89 \times \text{Weight} + 16.14 \times \text{Height} - 2.04 \times \text{Age} - 2244.45$ |
| **Kawasaki** | **Second morning urine** | $23 \times 16.3 \times (\text{Na spot} / \text{Crspot} \times \text{PrUCr24h})^{0.5}$<br>$\text{PrUCr24h} = 15.12 \times \text{Weight} + 7.39 \times \text{Height} - 12.63 \times \text{Age} - 79.9$ **(Male)**<br>$\text{PrUCr24h} = 8.58 \times \text{Weight} + 5.09 \times \text{Height} - 4.72 \times \text{Age} - 74.95$ **(Female)** |

## Results

Out of 120 participants, 92 (77%) were male and 28 (23%) were females. The mean BMI (Body mass index) was 39 (±11.47) and mean age was 26.5 (±5.0) years. The mean volume of 24 h urine was 1501 (±884) ml/day. Similarly, mean serum creatinine level was 0.88 (±.15)mg/dl ranged between 0.5 to 1.2 mg/dl and 24-hour urine creatinine clearance was 107.9 (±38.8) ml/min indicating normal kidney function. Mean levels of potassium, chloride, sodium,, and creatinine in spot and 24 hour urine are given in table 2. There was no significant difference between male and female for these analytes except sodium which was higher in females as compared to males (p.<0.002)

**Table 2:** Demographic Characteristics and Mean Levels Analytes.

| Description | Male<br>(n=92) | Female<br>(n=28) | Total<br>(n=106) | p-Value |
| --- | --- | --- | --- | --- |
| **Demography** |  |  |  |  |
| Age(years) | 39(±11.0) | 39(±12.0) | 39(±11.0) | 0.955 |
| Height(Feet) | 5.48(±0.39) | 5.46(±0.31) | 5.48(±0.37) | 0.749 |
| Weight(Kg) | 74.84(±14.30) | 71.25(±16.74) | 74(±14.91) | 0.311 |
| BMI(kg/m<sup>2</sup>) | 26.6(±5.2) | 26.6(±5.0) | 26.6(±5.1) | 0.946 |
| **Spot Urine** |  |  |  |  |
| Urine Creatinine(mmol/L) | 171(±104) | 133(±89.0) | 165(±102.0) | 0.139 |
| Spot Sodium (mmol/L) | 140(±64) | 115(±62.0) | 134(±64.0) | 0.081 |
| Spot Potassium (mmol/L) | 53.8(±33.6) | 51.6(±21.5) | 53.3(±33.3) | 0.693 |
| Spot Chloride (mmol/L) | 171(±78) | 132(±79.0) | 162±(79.0) | 0.027 |
| **24 h Urine** |  |  |  |  |
| Urine Creatinine (mmol/L) | 117.02(±58.4) | 102.39(±80.35) | 113.80(±63.78) | 0.393 |
| 24 hrs Sodium (mmol/L) | 158(±78) | 112(±60.0) | 148±(76.0) | 0.002 |
| 24 hrs Potassium (mmol/L) | 40(±17) | 46(±38.0) | 41±(23.0) | 0.421 |
| 24 hrs Chloride (mmol/L) | 148(±72) | 116(±71.0) | 141±(73.0) | 0.045 |

Mean 24 h sodium concentration was 158 mmol/l, which is almost equal to 8.64 ±4.43 g salt per day. There was a significant association between discretionary salt use with male gender (p. <0.004) and adding salt during cooking (p. <0.0001). However, there was no difference for age, BMI, adding salt on dinning, took salty food from restaurant, and knowledge about the impact of high salt on health (Table 3).

**Table 3:**
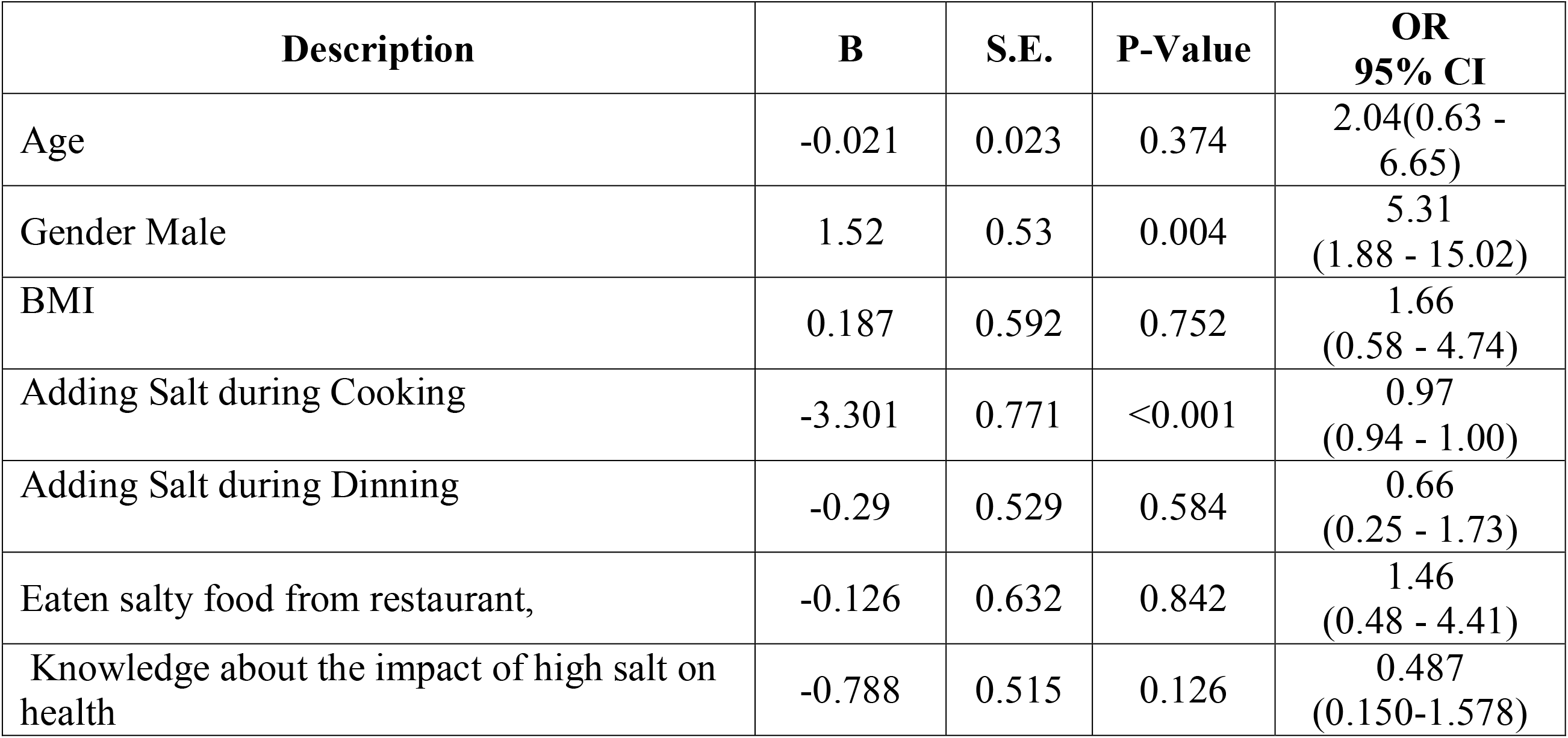
Association of discretionary salt with different risk factors.

A majority (71%) of the participants were using salt in their daily food while more than half were also adding extra salt at the dining table. When inquired from participants that how they categorized themselves regarding salt consumption, 91% believed it as per their need. Most (73%) of them were aware about the of the impact of extra use of salt on their health. Among all, 71% knew that it might cause hypertension and 11% associate this with bone diseases. Only few participants were aware that high level of salt might also cause gastric cancer and kidney stones. Twenty-five per cent of participants believed that reduction in daily salt intake is important for their health while others considered this either less important (57%) or gave no importance (17%).

Tanaka and Kawasaki equations showed significant difference between measured and estimated 24 hour sodium. However, the difference between measured and estimated 24 h sodium using INTERSALT method was insignificant (p. 0.09). Correlation analysis showed that none of three methods i.e. INTERSALT (bias: -19.64; CCC -0.79), Tanaka (bias: 167.35; CCC -0.37) and Kawasaki (bias: -42.49, CCC -0.79) showed agreement between estimated and measured 24 hour sodium. (Table 4 and Figure 1). Similarly, the Bland Altman plot also indicated an inconsistent pattern between estimated and measured 24 hour sodium (Figure 2).

**Table 4:** Comparison of measured (24 h) and estimated sodium.

| Description | Mean Difference | Stand Error | Concordance Correlation Coefficient | Intraclass Correlation Coefficient |
| --- | --- | --- | --- | --- |
| **Measured Na**(mmol/L) | Reference | Reference | Reference | Reference |
| **Intersalt**(mmol/L) | -19.64 | 11.65 | -0.79(-0.84 to -0.69) | -0.63(-0.74 to -0.50) |
| **Tanaka**(mmol/L) | 167.35* | 12.31 | -0.37(-0.53 to -0.19) | -0.33(-0.49 to -0.15) |
| **Kawasaki**(mmol/L) | -42.49* | 11.16 | -0.79(-0.85 to -0.70) | -0.67(-0.76 to -0.55) |

\*p. <0.05

**Figure 1:**
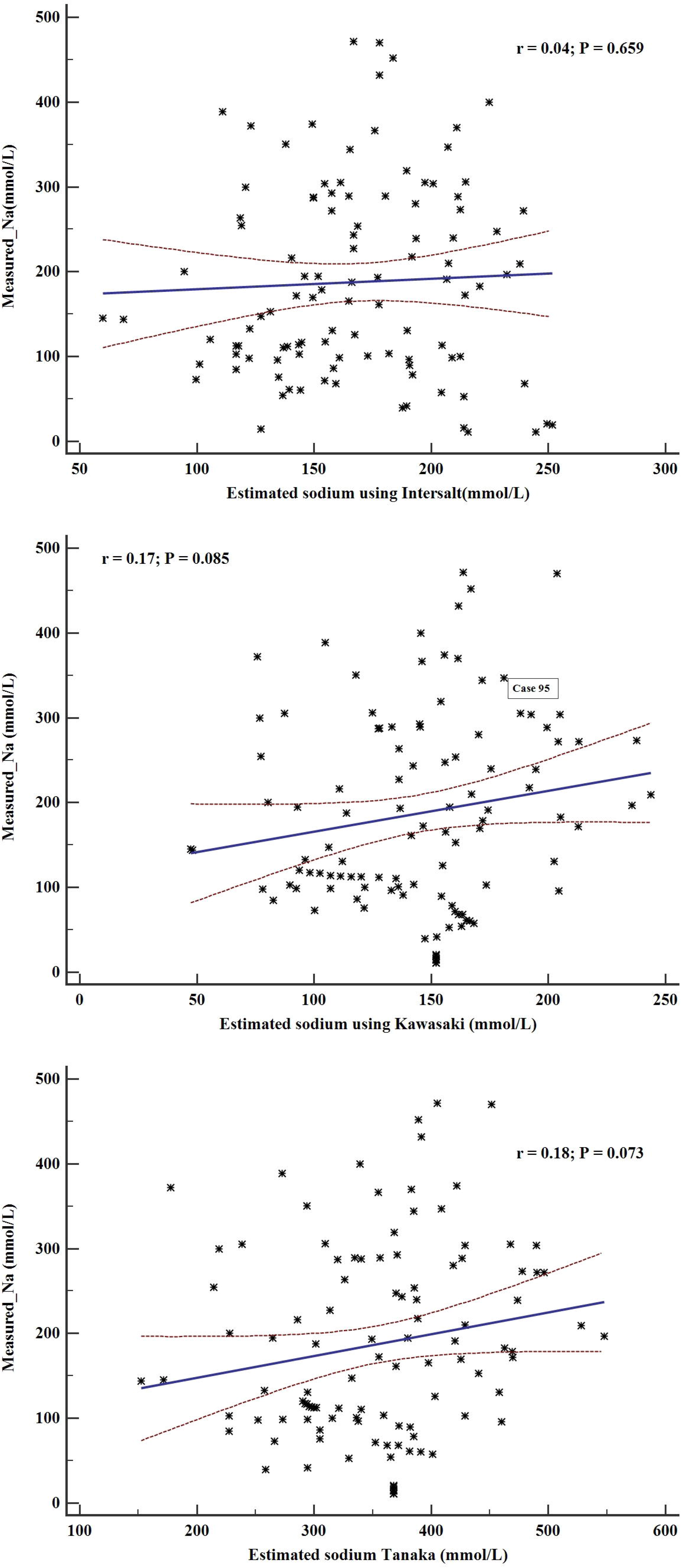
Scatter plot of measured 24 h urine sodium versus the estimated using Tanka (A), Kawasaki (B) and INTERSALT (C) methods. The black line is representing linear regression line while red dash lines are showing 95% predicted mean.

**Figure 2:**
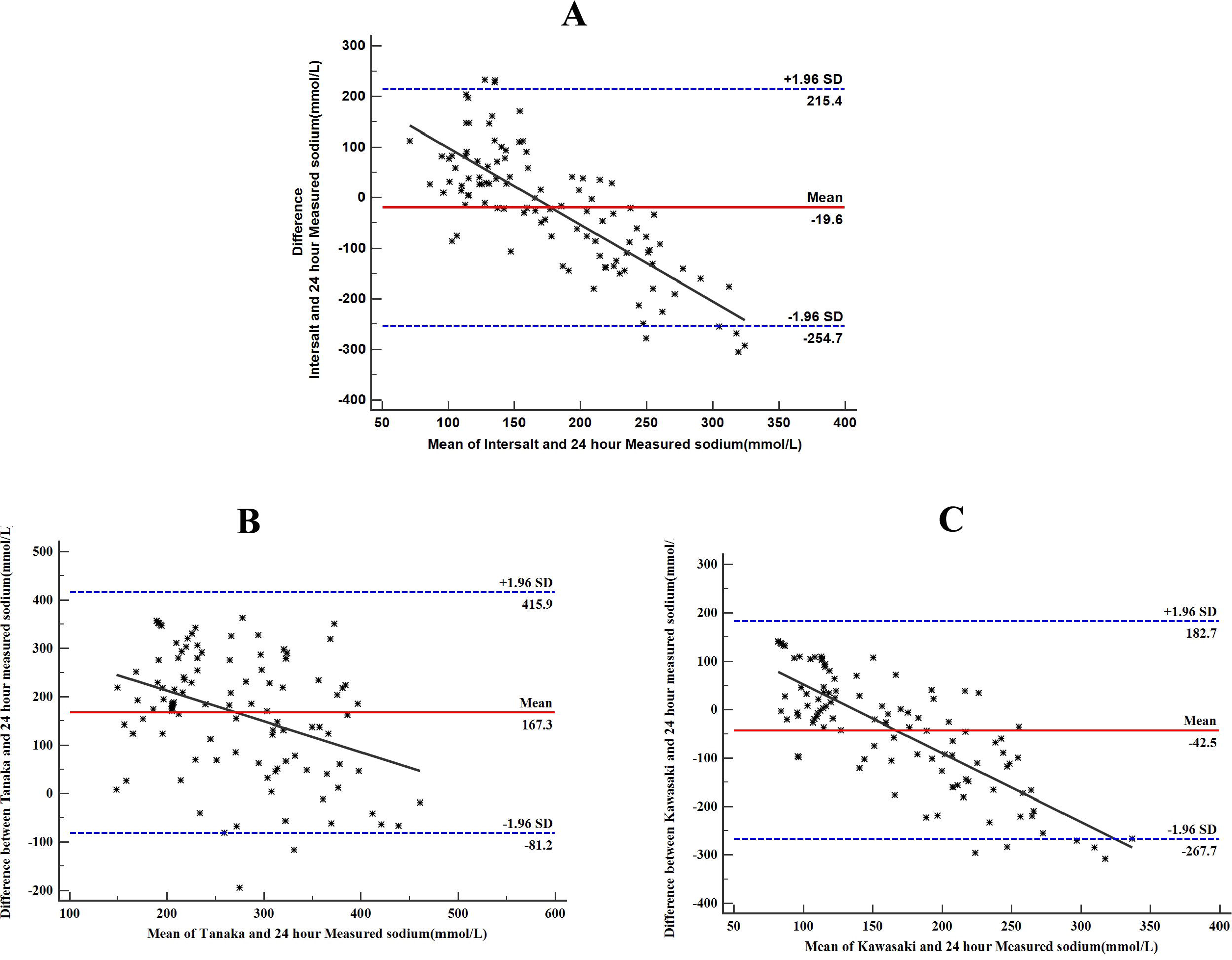
Bland Altman plot of measured 24-h urine sodium excretion versus INTERSALT (A), Kawasaki (B) and Tanaka (C) method estimated 24-h (mmol/L).The difference between measured and estimated was all estimated values minus the measured values. The mid red line is representing the mean difference or bias between measured and estimated values while the dashed line represents 95% limits of agreement of the mean difference (±1.96SD)

## Discussion

In this study, mean salt intake was 8.4 grams per day and was significantly associated with male gender and adding extra salt during dining at the table. The WHO recommends that salt intake should be less 5 grams per day(20). High salt intake has been associated with different diseases including hypertension, gastric cancer, kidney stones and bone deformities(21). Similar studies from India, Bangladesh and China also reported high salt intake of 10.98, 17 and 11.8 grams per day, respectively(22–24). High daily salt intake is also common in developed countries like in Australia where the mean daily intake of salt was 8.8 grams/day(25).

In Southeast Asia, excess salt intake is part of culinary culture, which results in increased consumption of daily salt in this region (26). However, the habit of adding salt during dinning is dangerous. Our study showed an association of discretionary salt use with male gender which is consistent with previous studies(27, 28). It was reported that high intake of salt in men might be due to high intake of food as compared to women (29). Another reason might be outside eating which more in men as compared to women in Pakistan.

Keeping in view the impact of high salt intake, the WHO members states agreed to reduce this by a relative of 30% by 2025(20). Many countries especially the developed ones have started different initiatives to reduce the daily salt intake. It has been shown that initiatives taken by the governments for the population-level reduction of salt intake were more useful (30). Pakistan is facing the dual burden of non-communicable and communicable diseases(31). It was reported that NCDs risk factors like tobacco, physical inactivity and unhealthy diet were common and there was a gradual increase in the burden of different NCDs especially diabetes, hypertension, and cardiovascular diseases (17, 32–35). The current findings of high daily salt intake is an addition to existing information on prevalence of different NCDs risk factors. Being the signatory of WHO treaty to lower the intake of salt in population, the Government of Pakistan must take urgent measures at policy level for reducing salt intake in the population. This will eventually help in reducing the burden of NCDs.

Estimating the salt intake (daily) is important to estimate the usage and monitor the impact of different interventions. The 24 hour urine is the standard method for estimating the intake of salt measuring the excretion of sodium in the urine. However, this is long and laborious and is impractical at population level. To overcome this issue, INTERSALT, Tanaka and Kawasaki methods (10, 15, 19) were formulated on concentrations of spot urine potassium, sodium, and creatinine along with gender, height, weight, and age.

The INTERSALT equation was derived using data of spot urine sodium concentration and 24 hour sodium excretion urine which was collected from 52 different populations belonging to 32 countries (19). On the other hand, both Kawasaki and Tanaka methods were derived using data of Japanese population(10, 15). Different studies have validated these methods to estimate daily salt intake using spot urine but their utility varied from population to population (13, 14, 16, 36–38). Additionally, these methods are inadequate to measure the intake of salt (daily) salt intake using spot urine sample but still excellent option for estimating at the population level(14).

The applicability of INTERSALT for estimating intake of salt was reported from different Western and Asian countries (30). It was shown that INTERSALT could be useful for estimating intake of salt by spot urine in the Asian population (12). In our study, the difference between estimated and measured sodium using INTERSALT (19) method was insignificant but the correlation analysis did not show any agreement, therefore, we do not support this notion. On the other hand, both Kawasaki and Tanaka methods had a significant difference between the means of estimated and measured 24 hour sodium. The correlation analysis also showed disagreement between estimated and measured 24 hour sodium excretion for both methods. Our study findings are consistent to previous reports indicating that probably these methods could not be used for all populations and there is need to make local equations(14, 39).

The value of creatinine concentration in a spot urine sample is important in predicting daily salt intake. Creatinine is produced during metabolization of creatinine phosphate in muscles. Hence its excretion in urine is dependent on body mass and protein intake. Similarly, creatinine excretion also varies among individuals depending on gender, age groups and other demogarphic features. The sodium to creatinine ratio might not depend on differnce at population level as there is low protein intake in developing countries (40) Also, Kawasaki and Tanaka’s methods derived using data of the Japanese population might not truly representing our population.

In summary, our study showed that salt intake (daily) is high though the people were aware of its impact on their health. The analysis also showed that the three methods i.e. Tanaka, Kawasaki and INTERSALT for 24 hour estimation are not applicable for Pakistani population. Our study provided a base for conducting large scale studies to formulate accurate equation for estimation of daily salt intake for local population. Besides this, interventional studies are also required for evaluating the possible interventions for behavior change in the population.

Our study has potential limitations like having less samples, lack of diversity with respect to ethnicity but still provides the valuable information regarding the validity of three methods for the estimating salt intake.

## Conclusion

Daily intake of salt was high than recommended by the WHO. Findings showed that none of the three methods could be used for estimating daily intake of salt in local settings of Pakistan.

## Data Availability

The data will be available.

## Authors Declaration

The MANS conceptualize and designed the study, secure the funding and revise the manuscript, IR drafted the project and manuscript, MA interpreted the data and revise the article with intellectual content, TR performed the statistical analysis and interpreted results.

## Ethics Statement

Institutional Bioethics Committee of Pakistan Health Research Committee provided ethical clearance. The research work carried according to the Helsinki Declaration and all participants gave written informed consent for their participation.

## Acknowledgments

We are thankful to the Excel labs staff for their dedication to test the samples timely especially Mr. Abdul Waheed, Manager Operations for his continuous support through the project. We are also obliged to all participants who voluntarily provided samples especially 24 hour urine sample. We are highly thankful to Mr. Saeed Ahmed Shahid for coordinating with the participants for collection of samples.

